# Sex-specific Morphometric Analysis of Ascending Aorta and Aortic Arch for Planning Thoracic Endovascular Aortic Repair

**DOI:** 10.1101/2023.02.22.23285632

**Authors:** Maria Katsarou, Tim J. Mandigers, Marton Berczeli, M. Mujeeb Zubair, Viony M. Belvroy, Adeline Schwein, Daniele Bissacco, Joost A van Herwaarden, Santi Trimarchi, Jean Bismuth

## Abstract

**Background:** In many studies on aortic disease women are underrepresented. The present study aims to assess sex-specific morphometric differences and gain more insight into endovascular treatment of the AA and arch.

**Methods:** Electrocardiogram-gated cardiac computed tomography scans of 116 patients who were evaluated for transcatheter aortic valve replacement were retrospectively reviewed. Measurements of the AA and aortic arch were made in multiplanar views, perpendicular to the semi-automatic centerline. Multiple linear regression analysis was performed to identify predictors affecting AA and aortic arch diameter in men and women. Propensity score matching was used to investigate whether sex influences aortic morphology.

**Results:** In both sexes, body surface area (BSA) was identified as a positive predictor and diabetes as a negative predictor for aortic diameters. In men, age was identified as a positive predictor and smoking as a negative predictor for aortic diameters. Propensity score matching identified 40 pairs. Systolic and diastolic mean diameters and AA length were significantly wider in men. On average, male aortas were 7,4% wider compared to female aortas, both in systole and diastole.

**Conclusions:** The present analysis demonstrates that, in women, increased BSA is associated with increased aortic arch diameters, while diabetes is associated to decreased AA and arch diameters. In men, increased body surface area and age are associated to increased AA and arch diameters, while smoking and diabetes are associated to decreased AA and arch diameters. Men were confirmed to have 7.4% greater AA and arch diameters than women.

## Introduction

Aneurysmatic disease of the ascending aorta (AA) and aortic arch is a potentially lethal but treatable condition. In the current era of endovascular aortic repairs, accurate assessment of aortic size is crucial for diagnosis, treatment and follow up. Endovascular repair of the AA is the final frontier in aortic surgery. The first ascending thoracic endovascular aortic repair (aTEVAR) for type A aortic dissection (TAAD) was reported in 2000 (1)(2)(3). Nowadays, it is more commonly employed in expert aortic centers, mostly to treat patients otherwise not fit for open surgical repair (4,5). Endovascular repair of the AA and arch is a valuable alternative to open surgery, providing acceptable early and mid-term outcomes (4) in patients who would otherwise face mortality rates of up to 95% when left untreated (6).

Aortic disease, being largely associated with atherosclerosis, has been linked to the male sex. However, more women are being treated nowadays due to an increasing aging population, and a change of social habits such as smoking, making it increasingly important to understand how sex differences might impact disease pathophysiology, prognosis, and treatment. Women have been traditionally underrepresented in many of the landmark studies which form the basis for guidelines recommendations, but contemporary research is increasingly focusing on sex-specific differences in aortic disease (7–9). The aim of the present study was to assess morphometric differences and identify different variables that might be associated with increased aortic size in the AA and aortic arch segments in the two sexes.

## Materials and Methods

### Study population

The Institutional Review Board (IRB) reviewed and waived ethical approval for this manuscript. A hundred and sixteen (116) consecutive patients who underwent Trans-catheter Aortic Valve Replacement (TAVR) at our Institution between September 2016 and February 2017 and had a preoperative, electrocardiography gated computed tomography angiography (ECG-gated CTA) scans were selected for morphometric analysis. Patients with aortic dissection, TEVAR, left ventricular ejection fraction <40% and those without ECG-gated CTAs were excluded.

### Data collection

Demographic data were collected for each patient including sex, age, race, body mass index (BMI), body surface area (BSA), smoking habits, aortic gradient, left ventricular ejection fraction and aortic arch type with retrospective chart review. Comorbidities including chronic kidney disease, diabetes mellitus, hypertension, coronary artery disease, cardiac heart failure, atrial fibrillation, chronic obstructive pulmonary disease, hyperlipidemia, connective tissue disease (CTD), history of coronary interventions were also assessed. History of aortic disease (arteritis and aneurysm), and prior aortic surgery were investigated as well.

### Measurement protocol

The initial 10 measurements were taken by two individual vascular surgeons (M.Z, A.S.) according to a previously published protocol (10), to ensure inter and intra-observer consistency. The rest were measured by a single vascular surgeon (M.Z.). Measurements were taken on multiplanar views perpendicular to a semi-automatically created aortic centerline on a single post-processing software workstation (Syngo.Via, Siemens Healthcare GmbH, Germany). R-R interval between 30-40% and 70-80% dictated the systolic and diastolic phases respectively. Inner aortic wall diameters were measured at the sinotubular junction (STJ), mid ascending aorta (at 4 centimeters proximal to the innominate artery (IA) ostium), proximally to the innominate artery, left common carotid (referred to as Ishimaru zone 1) and left subclavian (referred to as Ishimaru zone 2) ostium levels. Total ascending aortic length was measured from the STJ to the IA ostium.

Circumferential and arterial strain at each measuring point were calculated using the following equations:

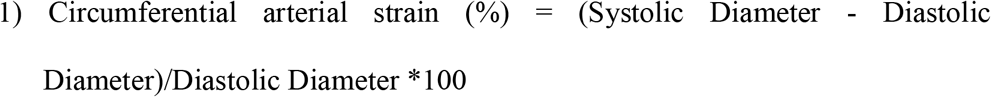

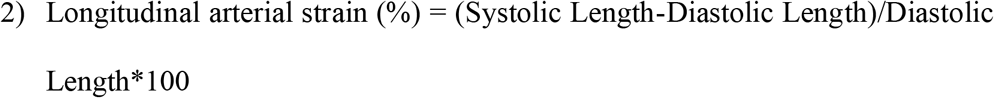

### Statistical analysis

The primary outcome of this study was to identify sex-specific variables associated with AA and arch size. The secondary outcome was to determine sex-specific morphometric differences and provide baseline measurements.

Multiple linear regression analysis was performed to identify variables potentially affecting ascending aortic and arch diameter in the male and female sex. The analysis was performed on the total initial cohort of patients. Age, BSA, smoking, diabetes and hypertension were chosen as potential clinical predictors for aortic diameter. In particular, BSA, age and diabetes have been previously identified as predictors for ascending aortic size (11). BSA has been identified to have a stronger correlation to aortic size than BMI (12) and therefore it was chosen as the most relevant variable to assess body size.

To mitigate sex-related biases, propensity score matching techniques were employed. Variables including age, BSA, BMI, CTD, hypertension, coronary artery disease, chronic heart failure, aortic arch type, aortic gradient, history of aortic disease and history of aortic surgery were utilized in the matching processes. A logistic regression was then performed to achieve similar baseline characteristics among the two groups using a 1:1 nearest neighbor matching technique with a 0.2 standard deviation caliper.

Numeric variables are expressed as means with standard deviation and compared through two sample t-test or Mann-Whitney U test. Nominal variables are expressed as number and percentages and compared with chi-square test or Fisher’s exact test.

In all analyses, p values < .050 were considered statistically significant. Statistical analysis was conducted using IBM SPSS 28 (Chicago, IL, USA).

## Results

### Inter and intra-observer variability

Inter and intra-observer variability was reported in our previous work (2); Intra-class correlation coefficient (ICC) and Bland-Altman analysis were performed. Inter-observer analysis showed good correlation for aortic diameter (ICC 0.99, mean difference -0.001 ± 0.52 mm) and aortic length (ICC 0.99, mean difference -0.03 ± 0.62 mm). Inter-observer analysis showed good correlation as well for aortic size (ICC 0.97, mean difference 0.14 ± 1.08 mm) and aortic length (ICC 0.99, mean difference -0.21 ± 3.01 mm).

### Baseline characteristics

Baseline demographics and comorbidities before and after propensity score matching are reported in table 1. Significant differences in demographics before matching were seen in race, BSA, smoking habits, and left ventricular ejection fraction. Our propensity score analysis yielded 40 matched pairs. The patients were well matched for age, race, BMI, smoking habits, aortic gradient and aortic arch type (normal or bovine). Significant differences were observed in BSA only (1.71 ± .21 vs 2.05 ± .16, p = .02). The two groups were well matched also for all comorbidities (Supplemental table S1).

**Table 1.** Baseline demographic characteristics.

|  | Total cohort |  |  | 1:1 ratio |  |  |
| --- | --- | --- | --- | --- | --- | --- |
|  | Women<br>(n=55) | Men<br>(n=61) | p | Women<br>(n=40) | Men<br>(n=40) | p |
| Age | 77.31±11.8<br>3 | 77.46±9.71 | .71 | 77.97±11.26 | 76.68±10.41 | .49 |
| Race |  |  | .01 |  |  | 1.0 |
| *Caucasian* | 36 (65.5) | 53 (86.9) |  | 26 (65) | 34 (85) |  |
| *Black* | 8 (14.5) | 3 (4.9) |  | 5 (12.5) | 1 (2.5) |  |
| *Hispanic* | 11 (20) | 3 (4.9) |  | 9 (22.5) | 3 (7.5) |  |
| *Other* | 0 (0) | 2 (3.3) |  | 0 (0) | 2 (5) |  |
| BMI | 29.01±7.23 | 27.75±4.86 | .72 | 27.67±5.69 | 27.37±4.74 | .76 |
| BSA | 1.74±.21 | 2.03±.20 | <.001 | 1.71±.21 | 2.05±.16 | .02 |
| Smoking |  |  | .01 |  |  | .85 |
| *Never* | 38 (69.1) | 25 (41) |  | 24 (60) | 24 (60) |  |
| *Active* | 3 (5.5) | 5 (8.2) |  | 2 (5) | 3 (7.5) |  |
| *Ex-smoker* | 14 (25.5) | 31 (50.8) |  | 14 (35) | 13 (32.5) |  |
| LVEF (%) | 65.36±11.3<br>7 | 58.46±11.71 | .002 | 65.85±10.96 | 58.33±10.96 | .94 |
| Aortic<br>gradient | 41.6±18.25 | 40.82±12.93 | .855 | 43.83±20.18 | 40.53±13.94 | .77 |
| Arch type |  |  | .61 |  |  | .77 |
| *1* | 43 (78.2) | 50 (82) |  | 31 (77.5) | 33 (82.5) |  |
| *2* | 12 (21.8) | 11 (18) |  | 9 (22.5) | 7 (17.5) |  |

Data are presented as *n* (%).
BMI = body mass index, BSA = body surface area, LVEF = left ventricular ejection fraction.

### Multiple linear regression analysis

In men, multiple linear regression analysis identified BSA as a positive predictor for aortic diameter from mid AA to zone 2 (mag. 6.45, p = .004; mag. 4.22, p = .022; mag. 4.42, p = .026; mag. 5.69, p = .001) (Figure 1), and age as a positive predictor for zone 2 diameter (mag. 0.104, p = .005). The presence of diabetes mellitus was a negative predictor for diameter from the STJ to zone 1 (mag. -2.99, p = .002; mag. -2.54, p = .005; mag. -1.66, p = .023; mag. -1.62, p = .04), and smoking was a negative predictor in mid-AA (mag. -0.97, p = .033). In women, the presence of diabetes in the distal AA and zone 1 (mag. -2.75, p = .017; mag. – 2.18, p = .03) was a negative predictor for diameter and BSA in zone 2 (mag. 5.34, p = .01) was a positive predictor. The multiple linear regression analysis is presented in Table 2.

**Figure 1.**
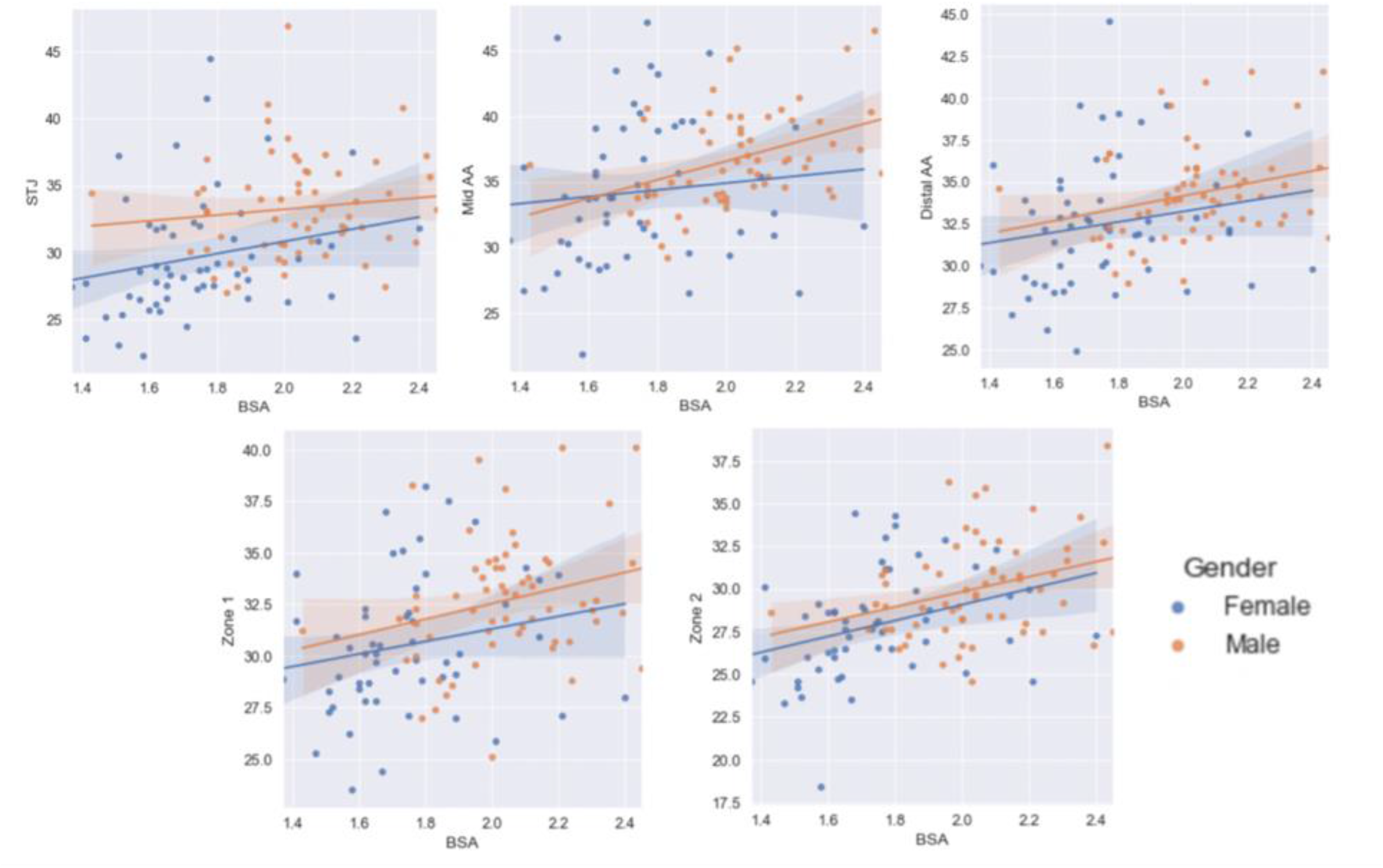
Linear regression scatter plots demonstrating the interaction between body surface area and aortic diameter (mm) for male and female sex. Male sex had a significant correlation with body surface area (m2) at mid and distal ascending aorta, zones 1 and 2 (p=.004, .022, .026, .001), while female sex showed correlation in zone 2 (p=.012) (STJ, SinoTubular Junction; AA, Ascending Aorta; BSA, Body Surface Area).

**Table 2.** Multiple linear regression analysis for predictors affecting ascending aortic size in men and women.

|  | STJ |  | Mid-AA |  | Distal AA |  | Zone 1 |  | Zone 2 |  |
| --- | --- | --- | --- | --- | --- | --- | --- | --- | --- | --- |
|  | Magnitude | p | Magnitude | p | Magnitude | p | Magnitude | p | Magnitude | p |
| **Men (n=61)** |  |  |  |  |  |  |  |  |  |  |
| Age | .004 | .939 | -.02 | .677 | .048 | .204 | -.052 | .207 | .104 | .005 |
| BSA | 1.65 | .484 | 6.45 | .004 | 4.22 | .022 | 4.42 | .026 | 5.69 | .001 |
| Smoking | -1.19 | .018 | -.97 | .033 | -.59 | .113 | -.49 | .227 | -.488 | .170 |
| DM | -2.99 | .002 | -2.54 | .004 | -1.66 | .023 | -1.62 | .040 | -.713 | .294 |
| HTN | 3.39 | .082 | 2.20 | .21 | -.57 | .696 | -1.20 | .449 | -2.56 | .068 |
| **Women (n=55)** |  |  |  |  |  |  |  |  |  |  |
| Age | -.07 | .181 | -.059 | .410 | -.02 | .670 | .005 | .893 | .016 | .685 |
| BSA | 3.84 | .207 | 2.63 | .490 | 4.10 | .100 | 4.06 | .064 | 5.34 | .012 |
| Smoking | -.48 | .505 | -.82 | .367 | -.46 | .429 | -.007 | .989 | -.17 | .718 |
| DM | -.80 | .561 | -1.77 | .307 | -2.75 | .017 | -2.18 | .030 | -1.00 | .238 |
| HTN | .21 | .916 | 1.42 | .573 | 1.67 | .309 | 2.48 | .088 | -.006 | .996 |

STJ = sinotubular junction, AA = ascending aorta, BSA = body surface area, DM = diabetes mellitus, HTN = hypertension.

### Differences in diameters and lengths and arterial strain

Differences in aortic measurements before and after propensity score matching are reported in Table 3. Considering the total cohort, before propensity score matching, systolic and diastolic diameters in the AA, aortic arch, and AA length were significantly different among women and men. In both men and women, the largest diameters were observed for the mid AA (36.81 ± 3.66 vs 34.23 ± 3.70, p = .004). All diameters progressively decreased when going distally from mid AA to zone 2. The percent difference in systolic diameters between men and women were 10.9% at the STJ, 7% in mid AA, 5.4% in distal AA, 6.5% in zone 1, 7% in zone 2, and 8.8% for AA length. Those differences remained similar for the diastolic measurements (10.4%, 6.9%, 5.8%, 6.1%, 7.9%, 9.2%, respectively). All diameters and AA length were greater in systole than in diastole. Diameters are illustrated in Figures 2 and 3.

**Table 3.** Ascending aorta and aortic arch measurements.

|  | **Total cohort** |  |  | **1:1 ratio** |  |  |
| --- | --- | --- | --- | --- | --- | --- |
|  | Women<br>(n=55) | Men<br>(n=61) | p | Women<br>(n=40) | Men<br>(n=40) | p |
| ***Systolic measurements*** |  |  |  |  |  |  |
| STJ | 29.66±4.55 | 33.3±3.79 | <.001 | 29.67 ± 4.74 | 33.92 ± 3.86 | .001 |
| Mid AA | 34.23±3.70 | 36.81±3.66 | .004 | 34.17 ± 5.76 | 37.48 ± 3.42 | .01 |
| Distal AA | 32.44±3.85 | 34.29±2.86 | <.001 | 32.78 ± 4.08 | 34.76 ± 2.85 | .03 |
| Length | 64.35±10.05 | 70.6±8.48 | <.001 | 64.86±10.42 | 71.60 ± 8.55 | .01 |
| Zone 1 | 30.52±3.33 | 32.66±3.06 | .001 | 30.97 ± 3.53 | 33.07± 2.97 | .004 |
| Zone 2 | 27.88±3.14 | 29.97±2.85 | .001 | 28.29 ± 3.34 | 30.41 ± 2.69 | .006 |
| ***Diastolic measurements*** |  |  |  |  |  |  |
| STJ diameter | 28.35±4.5 | 31.64±3.65 | <.001 | 28.29 ± 4.62 | 32.06 ± 3.78 | .001 |
| Mid AA | 33.49±5.53 | 35.99±3.44 | .002 | 33.29 ± 5.69 | 36.70 ± 3.27 | .008 |
| Distal AA | 31.56±3.61 | 33.52±2.84 | .001 | 31.84 ± 3.77 | 34.09 ± 2.93 | .012 |
| Length | 61.73±10.02 | 67.98±8.51 | <.001 | 62.18±10.38 | 69.16± 8.62 | .004 |
| Zone 1 | 29.92±3.34 | 31.85±2.99 | .001 | 30.26 ± 3.43 | 32.25 ± 3.01 | .016 |
| Zone 2 | 27.18±3.24 | 29.51±2.78 | .001 | 27.57 ± 3.37 | 29.81 ± 2.86 | .003 |
| ***Change between systole-diastole*** |  |  |  |  |  |  |
| STJ diameter | 1.30±.88 | 1.65±.95 | .049 | 1.38 ± .93 | 1.85 ± .97 | .042 |
| Mid AA | .80±.61 | .82±1.11 | .201 | .88 ± .62 | .78 ± 1.20 | .627 |
| Distal AA | .87±.95 | .76±.87 | .401 | .93 ± 1.06 | .67 ± .79 | .189 |
| Length | 2.61±1.94 | 2.61±1.91 | 3.66 | 2.67 ± 1.95 | 2.44 ± 1.70 | .542 |
| Zone 1 | .60±.71 | .80±.95 | .197 | .71 ± .65 | .82 ± .90 | .542 |
| Zone 2 | .70±.94 | .45±1.10 | .204 | .72 ± .97 | .60 ± .87 | .601 |
| ***Arterial strain (%)*** |  |  |  |  |  |  |
| STJ circ. | 4.72±3.28 | 5.29±3.14 | .954 | 4.96±3.46 | 5.89 ± 3.20 | .252 |
| Mid AA circ. | 2.46±2.07 | 2.33±3.16 | .280 | 2.71±2.10 | 2.16 ± 3.45 | .381 |
| Distal AA circ. | 2.77±2.76 | 2.33±2.61 | .124 | 2.92±3.02 | 2.02 ± 2.42 | .121 |
| AA longitudinal | 4.40±3.41 | 3.92±2.22 | .275 | 4.48±3.48 | 3.61±2.14 | .182 |
| Zone 1 circ. | 2.08±2.47 | 2.58±3.10 | .340 | 2.38±2.18 | 2.62±2.87 | .675 |
| Zone 2 circ. | 2.71±3.67 | 1.62±3.75 | .116 | 2.72±3.66 | 2.16±3.12 | .480 |

Data are presented as mean ± stand deviation (SD).
STJ = sinotubular junction, circ. = circumferential, AA = ascending aorta.

**Figure 2.**
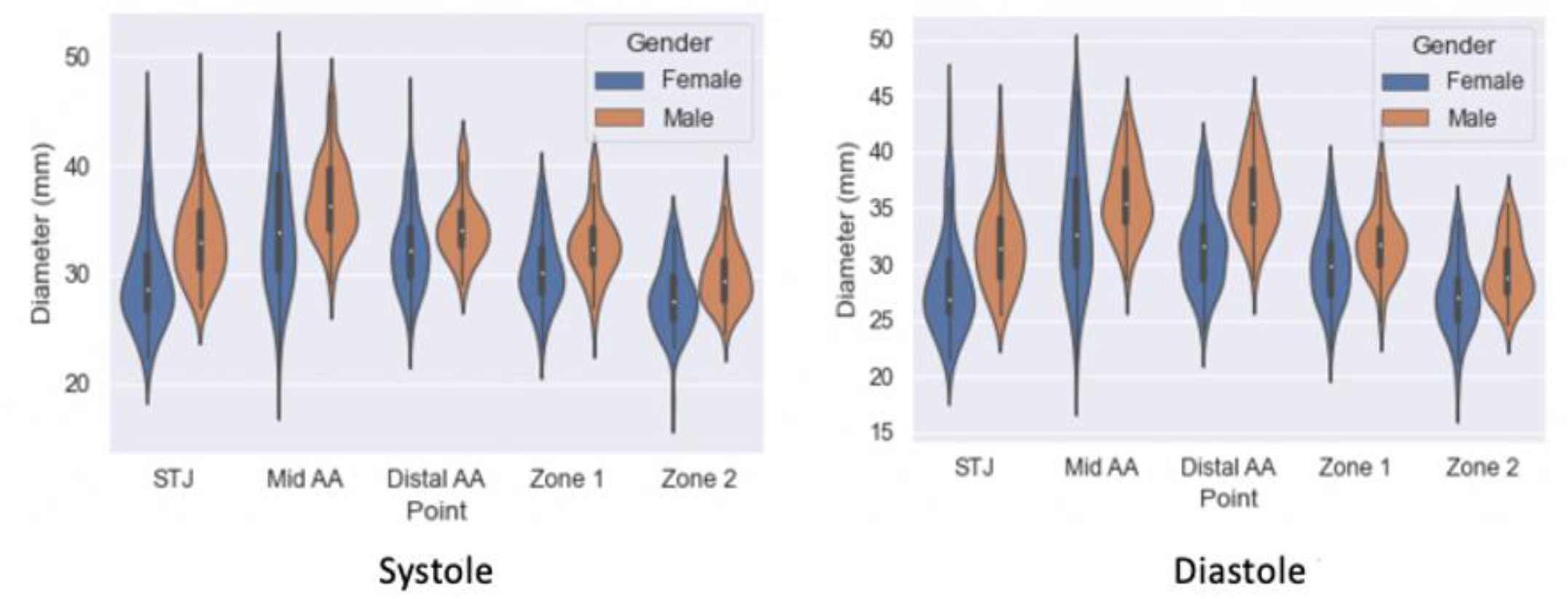
Violin plots comparing aortic diameters between women and men in systole and diastole. Mean diameter (white dot) in men is consistently larger across all the aortic points that were measured. Systolic diameters are higher than the diastolic counterpart in both genders. Largest diameters were measured at mid ascending aorta and smallest at zone 2.

**Figure 3.**
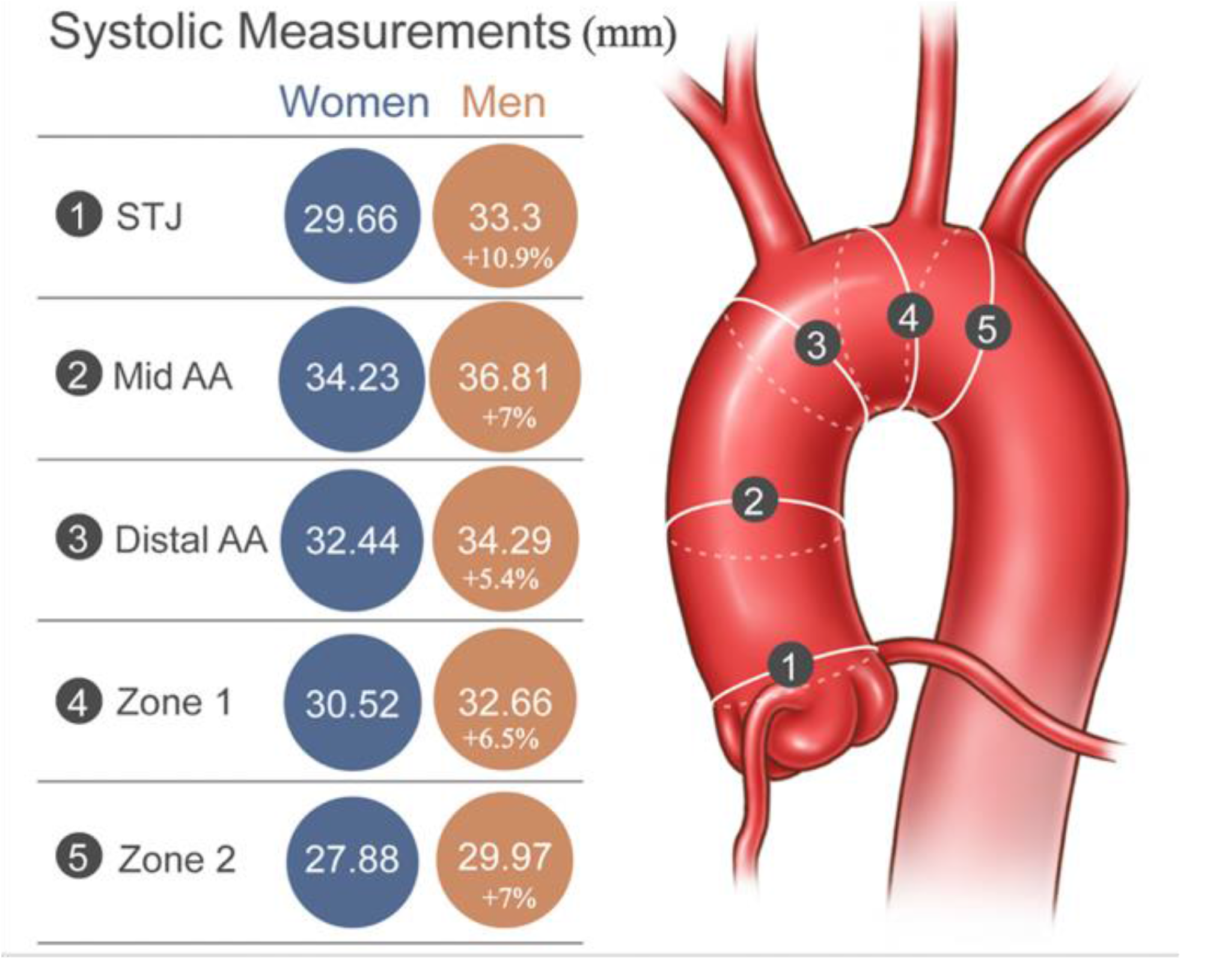
Graphical representation depicting the location where measurements were taken in the aorta, from sinotubular junction (STJ) to mid-ascending aorta (AA), distal AA, zones 1 and 2. On the left, cross-sections representing the diameter (mm) and % differences between men and women at each measurement point.

Arterial circumferential strain was more pronounced at the STJ (4.72 ± 3.28 for men vs 5.29 ± 3.14 for women, p = .95) and least pronounced at zone 1 for women (2.08 ± .2.47) and zone 2 for men (1.62 ± 3.75), progressively decreasing from proximally to distally. Longitudinal strain in the AA was 4.40 ± 3.41 vs 3.92 ± 2.2, p = .27. These values did not differ significantly between the two sexes.

When considering the group of matched patients, systolic and diastolic diameters and AA length also differed significantly at every point along the AA and arch. The percent difference in systolic diameters and AA length between the two sexes were 12.5% in the STJ, 8.8% in mid AA, 5.7% in distal AA, 6.3% in zone 1, 6.9% in zone 2, and 9.4% for AA length. On average, aortas in men were 8% larger. The differences for the diastolic measurements were 11.7%, 9.3%, 6.6%, 6.2%, 7.5%, 10% respectively. Arterial circumferential and longitudinal strain values did not differ significantly among the two sexes.

## Discussion

As women are increasingly being treated for aortic diseases, research focuses on sex-based differences for disease pathology, treatment, and outcome. Women are currently underrepresented in studies on TEVAR (13), especially aTEVAR (4,14,15), and have been shown to have more complications and worse surgical outcomes (7,9). In this study we attempted to gain more insight into AA and arch size and geometry between the two sexes. The demographic characteristics, comorbidities, and measurements of our cohort are consistent with the current literature (17–19). Our analysis provided baseline measurements for the AA in systole and diastole in both sexes, showing that diameters in the AA, aortic arch, and AA length were significantly different among women and men. On average, AA and aortic arch were 7,4% larger in men.

Our primary endpoint was to determine if there are any variables affecting AA and arch size in both men and women. In this context, multiple linear regression analysis revealed that increased age and BSA were positively associated with AA and arch diameters in men and BSA was positively associated arch diameters in women. Diabetes was negatively associated in both sexes, and smoking was negatively associated in men. Mori et al recently proposed a predictive model to identify patients with AA aneurysm (20), and they concluded that female sex and diabetes are associated with lower risk of AA aneurysm, whereas older age, higher BSA, hypertension and family history of aortic aneurysm were associated with an increased risk of an AA aneurysm. Wolak et al. also confirmed the association between BSA, diabetes and aortic size (11,12). The authors also suggest that male sex is a significant predictor only when interacting with age, meaning that older men have larger aortas than women of a similar age, but the difference is smaller for younger men and women. In our cohort, age was a positive predictor in zone 2 of men only, but this finding could be due to the decreased range of age among the patients evaluated. Smoking is traditionally considered as a risk factor for aneurysmal dilatation, but it was not found to be strongly correlated to aortic diameter. However, this could be inherent to the fact that ascending and descending aortic aneurysms have different etiopathogenic mechanisms.

Women generally have smaller and shorter arteries than men (18,19,21). With the advent of endovascular treatment of aortic arch pathologies, it is fair to question whether arterial size would impact endovascular treatment in women, or if they would need different endografts as compared to men. This should also be viewed in the light of the differences in diameter in zone 0 during the cardiac cycle, which might affect endograft size planning (22).

When considering the total cohort before propensity score matching, systolic and diastolic diameters and lengths were significantly different among both males and females in the AA. Men were found to have larger and longer AAs and arches. This finding is consistent with the work of Boufi et. al (19) who concluded that the mean difference in AA and arch diameter between men and women was 2.4 mm. In our cohort, the biggest difference was documented at the STJ with a mean difference of 3.5 mm, or 10.9%. Boufi also demonstrated that men have longer zone 0. We found that AA in men were, on average, 6 mm longer in both systole and diastole (8.8% and 9.19%, respectively). Interestingly, the change in length during the cardiac cycle was identical between the two sexes, averaging at 2.6 mm.

To mitigate potential bias related to sex, a propensity score matching was performed. Among the variables that we considered for the propensity score calculation was age, which has been shown to have a linear relationship with aortic measurements, mostly length (18,19,23). Rylski et al reported that women display a significantly greater increase in the size of the ascending and aortic arch segments with age than men (24). We also corrected for BSA which is also correlated to aortic morphology and has been shown to be more reliable than BMI for aortic dimensions (11). After correcting for BSA, Rylski et al. found that AA and aortic arch diameters were greater in women (24).

One of the main challenges with aTEVAR is correct sizing. Excessive oversizing might lead to aortic valve dysfunction or retrograde dissection while under sizing might lead to stent graft migration and possible flow disruption into the supra-aortic trunks. Even though the former holds true for most aortic endovascular interventions, it is even more important when treating the AA due to its natural hemodynamic and anatomical characteristics. Calculation of proximal landing zone diameter, most usually commonly at the STJ, is therefore essential when choosing a stent graft. ECG-gated CTA provides high resolution images and eliminates motion artifacts thus allowing for precise 3D measurements. In our cohort, change in STJ diameter between systole and diastole varied on average from 4.4% in women to 4.9% in men. Moreover, our mean population age was 77 years so we can assume that pulsatile changes might be even more pronounced in younger patients with less aortic stiffening (25). Csobay-Novak et al. (26) demonstrated that the largest diameter throughout the aorta is observed at 30% of the cardiac cycle. In our practice, the systolic phase at 40% of the cardiac cycle is used for planning measurements to avoid underestimation of oversizing.

ECG-gated CTA is the most common imaging modality used for studying the AA due to superior spatial resolution. However, several authors report morphometric data using alternative imaging modalities like trans-esophageal echocardiography (TEE) and magnetic resonance (MR) (27–29). MR lacks spatial resolution but offers the advantage of decreased radiation exposure. TEE allows for simultaneous functional cardiac evaluation, which can be useful in the preoperative setting. Rodriguez-Palomares et al. evaluated the AAs of 140 patients with TEE, CTA, and MR (29). The authors concluded that aortic root and AA diameters measured by TTE using the leading edge-to-leading edge convention showed accurate and reproducible values compared with internal diameters assessed by CTA or MR. The good correlation between the three most common modalities permits multi-modality follow-up of patients with aortic disease without any impact on aortic measurement accuracy.

### Limitations

One of the limitations of this study is the limited number of patients included in the study cohort and the retrospective, single-center nature of study design. Moreover, patients with aneurysms or dissections were excluded from the study so the values collected may not be representative in patients with those diseases. Data were drawn from a selected cohort of patients who underwent TAVR for aortic stenosis, which could introduce a selection bias. However, this study provides insight into healthy aortas, thus eligible proximal landing zones for endovascular procedures. Considering the older age of the cohort, conclusions might not be drawn for younger populations, but most aortic interventions interest older patients. Another limitation is that measurements were performed mostly by a single operator, but the intra and inter-observer variability cohort analysis identified small differences of under 1 mm. Small changes in size can be simply identified on ECG-gated CTA because of the high spatial resolution. However, out of plane aortic movement might have caused minimal miscalculations, that is inherent to imaging studies.

## Conclusions

Although future studies with larger sample sizes are needed to better understand sex-specific morphological variations and their potential impact on endovascular aortic repair, our findings are of importance, both for physicians and device manufacturers, for clarifying some of the current gaps in endovascular programs’ development of ascending aorta and arch. Using body surface indexed measurements may decrease sex-related anatomic disparities. Increased awareness and knowledge about sex-specific differences in aortic disease are important to improve patient outcomes and tailor endovascular procedures and materials to female needs.

## Data Availability

A dataset used to extrapolate statistical data is available.

## Acknowledgements

We thank Racheal E. Whitehead, scientific illustrator, for anatomical sketches used in our illustrations to describe the results.

